# Vaccine effectiveness against severe COVID-19 during the Omicron wave in Germany: Results from the COViK study

**DOI:** 10.1101/2023.01.09.23284327

**Authors:** Anna Stoliaroff-Pepin, Caroline Peine, Tim Herath, Johannes Lachmann, Wiebke Hellenbrand, Delphine Perriat, Achim Dörre, Andreas Nitsche, Janine Michel, Marica Grossegesse, Natalie Hofmann, Thomas Rinner, Claudia Kohl, Annika Brinkmann, Tanja Meyer, Daniel Stern, Fridolin Treindl, Brigitte G. Dorner, Sascha Hein, Laura Werel, Eberhard Hildt, Sven Gläser, Helmut Schühlen, Caroline Isner, Alexander Peric, Ammar Ghouzi, Annette Reichardt, Matthias Janneck, Guntram Lock, Dominik Huster, Thomas Grünewald, Lars Schaade, Ole Wichmann, Thomas Harder

**Affiliations:** Department for Infectious Disease Epidemiology, Immunization Unit, Robert Koch Institute; Department for Infectious Disease Epidemiology, Robert Koch Institute; Centre for Biological Threats and Special Pathogens, ZBS1 Highly Pathogenic Viruses, WHO Reference Laboratory for SARS-CoV-2 and WHO Collaborating Centre for Emerging Infections and Biological Threats, Robert Koch Institute; Centre for Biological Threats and Special Pathogens, ZBS3 Biological Toxins, Robert Koch Institute; Division Virology, Paul-Ehrlich-Institute; Klinik für Innere Medizin - Pneumologie und Infektiologie, Vivantes Klinikum Neukölln und Spandau, Berlin; Vivantes Netzwerk für Gesundheit GmbH, Direktorat Klinische Forschung & Akademische Lehre, Berlin; Klinik für Innere Medizin - Infektiologie, Vivantes Auguste-Viktoria-Klinikum, Rubensstr. 125, 12157, Berlin; Klinik für Pneumologie und Infektiologie, Vivantes Klinikum im Friedrichshain, Landsberger Allee 49, 10249 Berlin; Schön Klinik Düsseldorf, Interdisziplinäre Notaufnahme, Am Heerdter Krankenhaus 2, 40549 Düsseldorf; Helios Klinikum Berlin-Buch, Schwanebecker Chaussee 50, 13125 Berlin; Klinik für Kardiologie, Sektion Nephrologie, Albertinen Krankenhaus, Süntelstraße 11a, 22457 Hamburg; Klinik für Innere Medizin, Albertinen Krankenhaus, Süntelstraße 11a, 22457 Hamburg; Klinik für Innere Medizin, Helios Klinikum Erfurt; Klinik für Infektions- und Tropenmedizin, Klinikum Chemnitz; Centre for Biological Threats and Special Pathogens, Robert Koch Institute

**Keywords:** COVID-19, vaccine effectiveness, hospitalization, case-control study, SARS-CoV-2, Omicron

## Abstract

**Purpose:** COViK - a prospective hospital-based multicenter case-control study in Germany - aims to assess the effectiveness of COVID-19 vaccines against severe disease. Here, we report vaccine effectiveness (VE) against COVID-19-caused hospitalization and intensive care treatment during the Omicron wave.

**Methods:** We analyzed data from 276 cases with COVID-19 and 494 control patients recruited in 13 hospitals from 1 December 2021 to 5 September 2022. We calculated crude and confounder-adjusted VE estimates.

**Results:** 21% of cases (57/276) were not vaccinated, compared to 5% of controls (26/494; p < 0.001). Confounder-adjusted VE against COVID-19-caused hospitalization was 55.4% (95% CI: 12-78%), 81.5% (95% CI: 68-90%) and 95.6% (95%CI: 88-99%) after two, three and four vaccine doses, respectively. VE against hospitalization due to COVID-19 remained stable up to one year after three vaccine doses.

**Conclusion:** Three vaccine doses remained highly effective in preventing severe disease and this protection was sustained; a fourth dose further increased protection.

**Ethical review and trial registration:** The study was approved by the Ethics Committee of the Charité Universitätsmedizin, Berlin (EA1/063/21) and was registered at “Deutsches Register Klinischer Studien” (DRKS00025004).

**Potential conflicts of interest:** S. G. received payment/honoraria from Astra Zeneca, Boehringer Ingelheim, Roche Pharma, Novartis and Berlin Chemie, this had no influence on this work; all other authors reported no conflicts of interest.

## Background

The SARS-CoV-2 pandemic spread globally since December 2019 with more than 600 million cumulative cases and 6.4 million deaths documented by the end of August 2022 [1]. Several different variants of the virus evolved over time. In Germany, the Alpha and Delta variants dominated in 2021. In November 2021, the WHO declared Lineage B.1.1.529, also called Omicron variant, as a variant of concern (VOC). From 12 December 2021 to 30 January 2022, the proportion of the Omicron variant in Germany increased from 2.3% to 97.8% [2].

The Omicron variant differs from former variants by more than 30 amino acid modifications in the spike protein. It is characterized by many concerning epidemiological features like lower minimal infection dose resulting in higher transmissibility, immune evasion with the risk of reinfections and breakthrough infections, and an impaired response to COVID-19-specific treatment [3-5]. In contrast to other variants, virus replication of the Omicron variant occurs mainly in the upper respiratory tract (e. g. pharynx), which may lead to higher transmission rates and milder disease [5-8].

Vaccine effectiveness (VE) against the wild-type virus and the Alpha, Beta and Delta variants was very high irrespective of outcome definition [9, 10]. Previous studies showed decreased VE against the Omicron variant due to mutations in the spike protein, but effectiveness against severe disease remained high [11, 12]. Many study teams analyzing VE during the pandemic used registry data, which do not always allow differentiation between COVID-19 as the main diagnosis or SARS-CoV-2 infection as secondary diagnosis. This may lead to biased VE estimates due to incorrect classification of the endpoint.

More detailed, individual data are needed to allow valid conclusions about the VE in subgroups with proper statistical adjustment for confounders. We therefore launched a case-control study in 13 hospitals across Germany (COViK) to assess the effectiveness of vaccines in preventing COVID-19-associated hospitalization in the adult population. In addition, we performed subgroup analyses and studied different time intervals of protection. Initial data collected during the Delta wave were published recently [9]. Here we present the profile of hospitalized COVIK-19-patients and COVID-19 VE based on data obtained during the Omicron wave.

## Methods

Study design: We conducted a prospectively recruiting multi-center case-control study with 1:2 matching in 13 hospitals in five federal states (Berlin, Hamburg, North Rhine-Westphalia, Thuringia, Saxony) in Germany. For this interim analysis, we used data of patients recruited during the Omicron wave.

Study nurses were trained by the COViK study center team at the beginning of the assignment. Additionally, on-site audits were conducted regularly and study nurses underwent several in-house trainings.

Inclusion/ exclusion criteria and matching: Patients were eligible to be included, if they were aged between 18 and 90 years and hospitalized due to laboratory-confirmed COVID-19 at one of the 13 study hospitals from 1 December 2021 until 5 September 2022. Controls were SARS-CoV-2 negative patients in the same hospital (or a hospital in the same city) as the case and were matched (1:2) by age, sex and admission date (for details, see supplement).

Biological samples and data collection: Nasopharyngeal and oropharyngeal swabs were taken from cases and controls. SARS-CoV-2 PCR and sequencing were performed at the Robert Koch Institute [13, 14] (for details, see supplement). Information about socio-demographic factors, course of COVID-19, vaccination history, risk factors for COVID-19 infection and severe course of disease were collected during interviews. Clinical and laboratory data were extracted from medical records. We developed our questionnaire to permit estimation of the Charlson comorbidity index [15].

Study nurses validated the information on vaccination with the vaccination certificate or vaccination app.

### Statistical analysis

Comparisons between cases and controls were performed using appropriate significance tests (unpaired Student’s t-tests for age and BMI and Chi-squared test or Fisher’s exact test for age group, sex, educational level, number of comorbidities, vaccination status and number of vaccine doses).

We computed the 2-dose, 3-dose, 4-dose and 2 vs. 3 or 4 dose-VE for all patients, as well as for the following subgroups: males/females; age 18-59/60-69/70-90 years; <3/≥3 pre-existing comorbidities; admission to intensive care (ICU)/non-ICU ward. For assessment of the duration of vaccine-induced protection, VE was estimated according to symptom onset 14-90 days, 91-180 days and 181-365 days after the last vaccine dose. Patients who received only one vaccine dose were included in the descriptive analysis but excluded from VE analysis.

As the matched subgroup-analysis was not applicable for all subgroups due to insufficient number of patients in the strata, we primarily performed an unmatched analysis as also suggested by others [16]. The odds ratio (OR) to prevent severe COVID-19 by vaccination was calculated with the formula

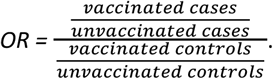

The VE was subsequently calculated with the formula

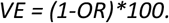

To assess the robustness of results, pairwise matched analysis was conducted (see Supplement Tables 3 and 4, Figure 4) using the Mantel-Haenszel method:

Matched pairs odds ratio: 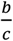

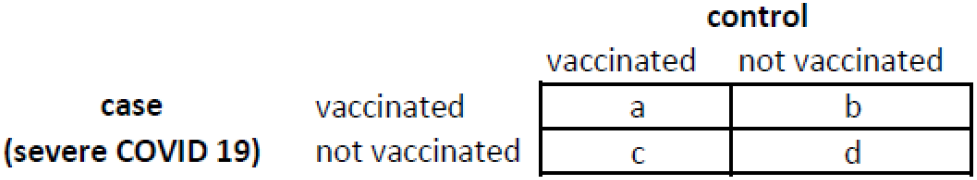

In some subgroup analyses, we applied the Woolf-Haldane correction as the combination vaccinated case(s)/unvaccinated control(s) was not present in every subset of the data.

Immunocompromising comorbidities/ therapies and diseases without immunosuppression were analyzed separately. To determine the minimal set of relevant confounder variables for the adjustment we constructed a directed acyclic graph (DAG; see Supplement Figure 2). Accordingly, estimation of VE was adjusted for age group, socio-economic status (education), pre-existing comorbidities and risk of infection proxies (e. g. profession, daily high-risk activities without masking, housing situation); see also “Supplementary Methods”). For four variables, adjustment was already performed through matching (sex, region, phase of pandemic, infection protection recommendations).

Data were analyzed using the statistical software R, version 4.1.2.

## Results

We recruited 770 participants, comprising 276 cases and 494 matched controls (Table 1) during the specified study period. Sociodemographic data: The median age of cases was 71 years and of controls, 68 years (Figure 1).

**Table 1.** Characteristics of cases and controls.

|  | Cases | Controls | p-value |
| --- | --- | --- | --- |
| **n** | 276 | 494 |  |
| **Age (years) mean (SD)** | 66.65 (16.18) | 65.25 (15.28) | 0.233 |
| **Age group n (%)** |  |  |  |
| 18-59 years | 78 (28.3) | 150 (30.4) | 0.355 |
| 60-69 years | 55 (19.9) | 114 (23.1) |  |
| 70-90 years | 143 (51.8) | 230 (46.6) |  |
| **Sex n (%)** |  |  |  |
| male | 155 (56.2) | 291 (58.9) | 0.506 |
| female | 121 (43.8) | 203 (41.1) |  |
| **Highest school educational level n (%)** |  |  |  |
| no graduation | 16 (5.8) | 16 (3.2) | 0.092 |
| 9 school years | 69 (25.0) | 113 (22.9) |  |
| 10 school years (secondary school certificate) | 128 (46.4) | 218 (44.1) |  |
| 12 or 13 school years (high school graduation) | 63 (22.8) | 147 (29.8) |  |
| **BMI (kg/m<sup>2</sup>) mean (SD))** | 26.93 (6.14) | 29.00 (18.11) | 0.066 |
| **BMI group n (%)** |  |  |  |
| underweight/normal weight (BMI ≤ 25 kg/m<sup>2</sup>) | 124 (44.9) | 188 (38.1) | 0.131 |
| overweight (BMI > 25-30 kg/m<sup>2</sup>) | 79 (28.6) | 171 (34.6) |  |
| obese (BMI > 30 kg/m<sup>2</sup>) | 73 (26.4) | 135 (27.3) |  |
| **Admission to intensive care unit (ICU) n (%)** |  |  |  |
| yes | 23 (8.3) | NA |  |
| no | 253 (91.7) | NA |  |
| **Death n (%)** |  |  |  |
| yes | 9 (3.3) | NA |  |
| no | 267 (96.7) | NA |  |
| **Pre-existing comorbidities (general) n (%)** |  |  |  |
| < 3 | 166 (60.1) | 350 (70.9) | 0.003 |
| ≥ 3 | 110 (39.9) | 144 (29.1) |  |
| **Pre-existing comorbidities (immune system) n (%)** |  |  |  |
| none | 191 (69.2) | 414 (83.8) | < 0.001 |
| ≥ 1 | 85 (30.8) | 80 (16.2) |  |
| **Vaccination (≥ 2 doses) n (%)** |  |  |  |
| yes | 206 (74.6) | 461 (93.3) | <0.001 |
| no | 70 (25.4) | 33 (6.7) |  |
| **Number of vaccine doses n (%)** |  |  |  |
| 0 | 57 (20.7) | 26 (5.3) | <0.001 |
| 1 | 13 (4.7) | 7 (1.4) |  |
| 2 | 46 (16.7) | 49 (9.9) |  |
| 3 | 145 (52.5) | 350 (70.9) |  |
| 4 | 15 (5.4) | 62 (12.6) |  |
| **Vaccine type n (%)** |  |  |  |
| one dose (mRNA or vector) | 13 (4.7) | 7 (1.4) |  |
| two doses (mRNA/mRNA) | 42 (15.2) | 41 (8.2) |  |
| two doses (vector/vector) | 1 (0.3) | 4 (0.8) |  |
| two doses( crossover<sup>b</sup>) | 2 (0.7) | 4 (0.8) |  |
| three doses (mRNA/mRNA/mRNA) | 126 (45.6) | 281 (56.8) |  |
| three doses (mRNA/mRNA/vector) | 0 | 1 (0.0) |  |
| three doses (vector/vector/mRNA) | 10 (3.6) | 29 (5.8) |  |
| three doses (crossover<sup>b</sup>/vector) | 1 (0.3) | 0 (0) |  |
| three doses (crossover<sup>b</sup>/mRNA) | 7 (2.5) | 39 (7.8) |  |
| not vaccinated | 57 (20.6) | 26 (5.2) |  |
| four doses, other vaccine or missing information | 17 (6.1) | 62 (12.5) |  |

<sup>a</sup> High-risk comorbidities for severe course of COVID-19: diabetes type 2, Organ transplant, BMI > 30, COPD, renal insufficiency, heart failure [17]
<sup>b</sup> first dose mRNA, second dose vector vaccine or vice versa

**Figure 1.**
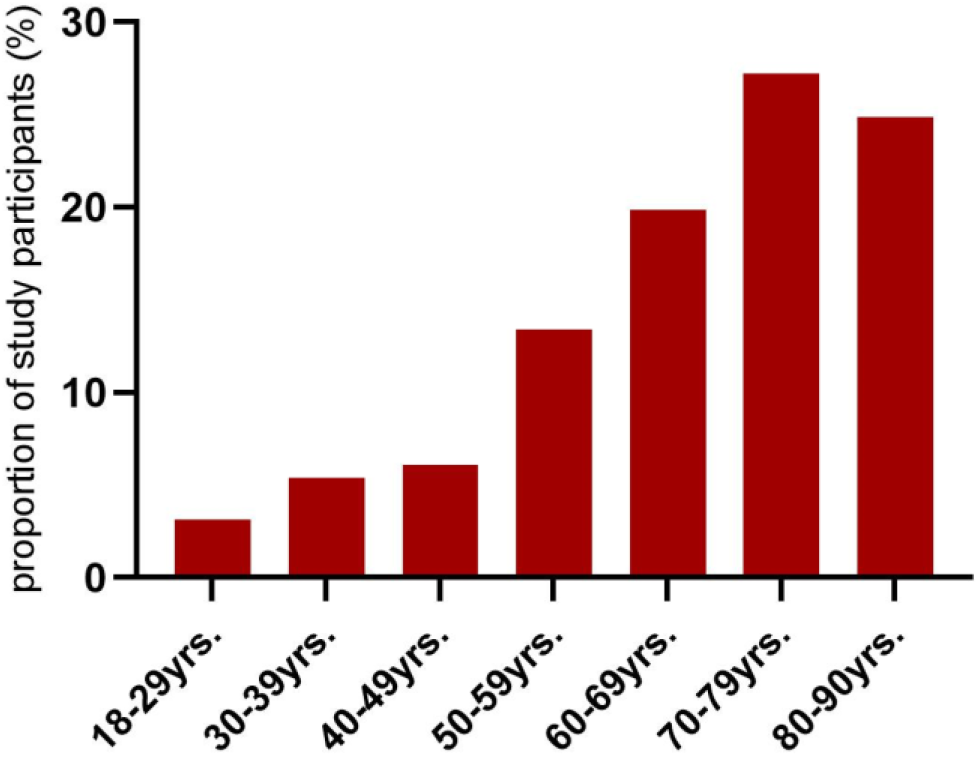
Age group proportions in the Omicron wave (cases)

Of cases, 44% were female and of controls, 41%. Cases and controls were similar regarding their level of education, with 31% of cases and 26% of controls reporting nine or fewer school years (p = 0.092, Table 1).

Immunocompromising comorbidities were more prevalent in cases (31%) than in controls (16%, p < 0.001, Figure 2, Table 1). Other comorbidities (e. g. heart failure, renal insufficiency, diabetes mellitus) were also more frequent in cases compared to controls (cases with ≥ 3 comorbidities 40%, controls 29%, p= 0.003, Figure 2, Table 1).

**Figure 2.**
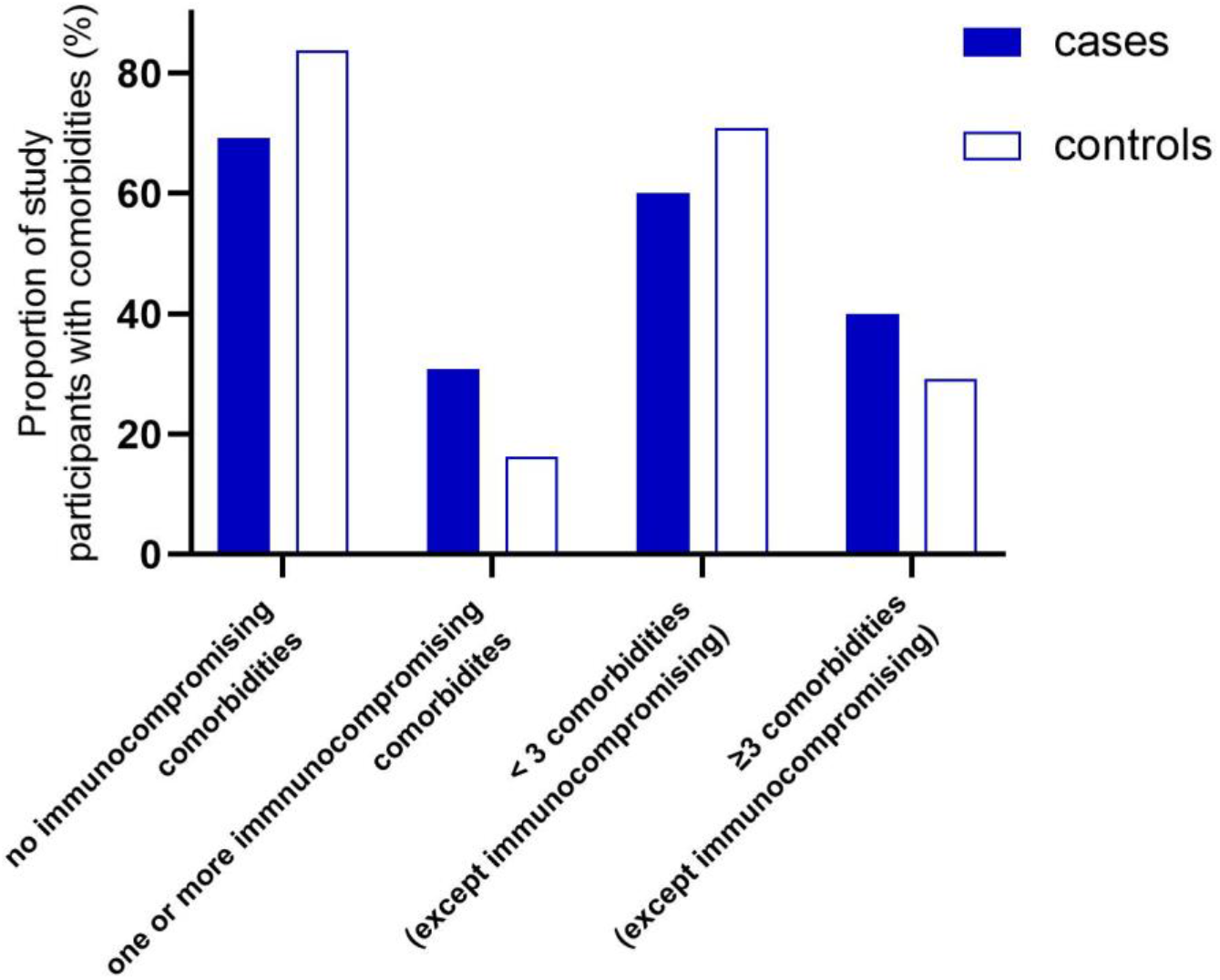
Comorbidities of study participants in the Omicron wave, cases vs. controls.

Intensive Care Unit (ICU) treatment was necessary for 23 cases (8.3%). Nine cases died during their hospital stay (3.3%, Table 1).

Subtyping showed most cases to be infected with Omicron belonging to the BA.1 and BA.2 subgroups (Supplement Figure 3).

Vaccination status differed significantly between cases and controls: 21% of the cases were not vaccinated at all, compared to 5% of the controls (p < 0.001, Figure 3). Moreover, only 53% of cases, compared to 71% of controls, had received three doses of vaccine. 5% of cases and 13% of controls had received four vaccine doses (Figure 3). The difference between the proportions of boostered (three or four doses) cases and controls was statistically significant (p < 0.001).

**Figure 3.**
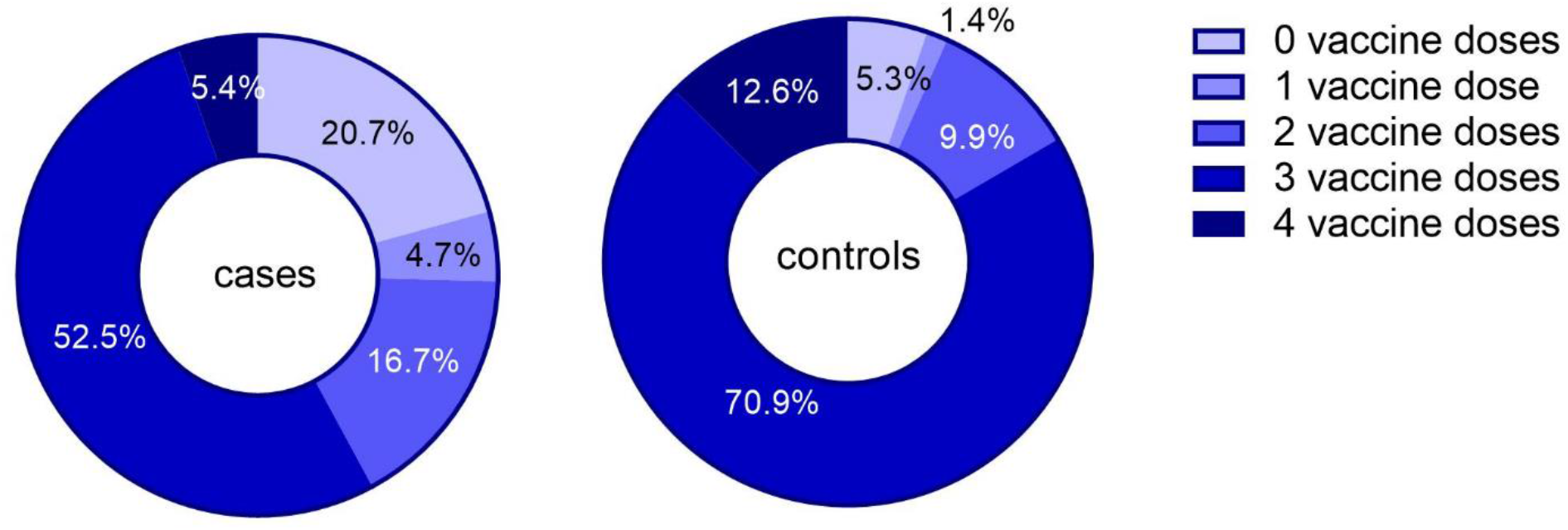
Number of vaccine doses in cases (n = 276) and controls (n = 494)

For 238 participants (35%), information on vaccination was verified by the vaccination certificate. In 374 participants (54%), the vaccination app was used for verification, and in 14 participants (2 %), this information came from the general practitioner. It was not possible to verify the data of 61 vaccinated participants (9%). 83 participants were not vaccinated. In the unmatched crude analysis, two-dose VE was 56.4% (95% CI: 19-76%) three-dose VE was 80.8% (95% CI: 68-88%) and 4-dose VE was 88.7% (95% CI: 76-94%). Compared to 2 doses, VE was 58% (95% CI: 35-73%) for ≥ 3 doses (Figure 4). For further details, see Supplement Tables 1 and 2).

**Figure 4.**
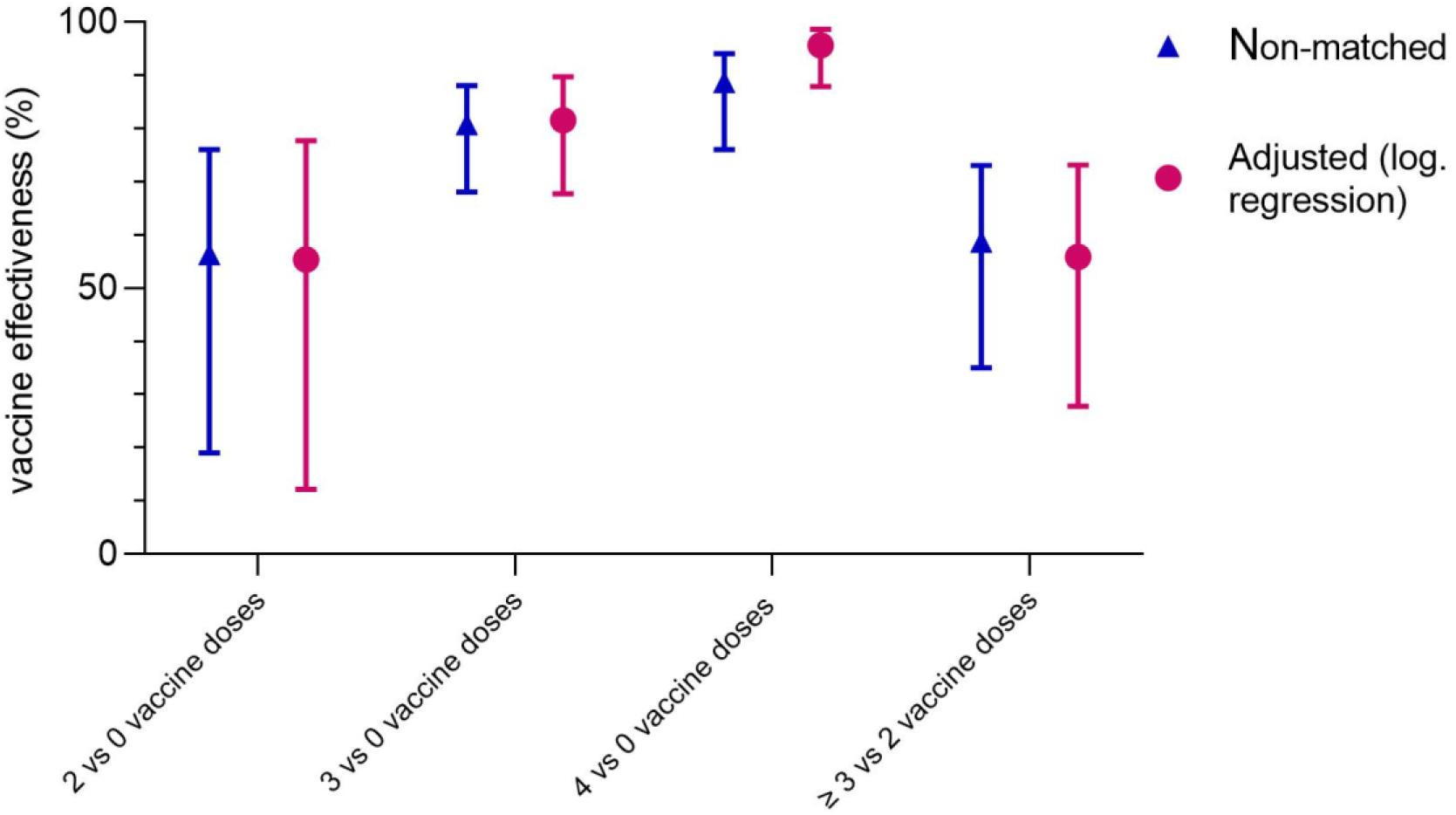
Vaccine effectiveness: unmatched analysis, crude versus confounder adjusted logistic regression.

After adjustment for confounders, overall VE in the Omicron wave was similar to the results of the crude analysis (Figure 4). The adjusted VE against severe disease was 55.4% (95% CI: 12-78%) after two, 81.5% (95% CI: 68-90%) after three and 95.6% (95% CI: 88-99%) after four doses compared to non-vaccinated.

The results of the pairwise matched analysis were similar to those of the unmatched analysis (see Supplement Tables 3 and 4, Supplement Figure 4).

VE after four doses was about 90% in all subgroups, with the highest estimate in ICU treated patients (96.5%; 95% CI: 35-99%), and in the age group > 70 years (94.9%, 95% CI: 85-98%). Four-dose VE estimates was higher (although not significantly) than 3-dose VE in all subgroups (Supplement Table 2).

When VE estimates were stratified according to time interval between last vaccine dose and date of symptom onset, we observed no significant differences between VE values, indicating a stable VE over time for up to one year. As shown in Figure 5, this was observed for two and three vaccine doses. Six to twelve months after the last vaccine dose, VE was 59.6% (95% CI: 13-81%) for two doses and 81% for three vaccine doses (95% CI: 63-90%, Supplement Tables 1 and 2).

**Figure 5.**
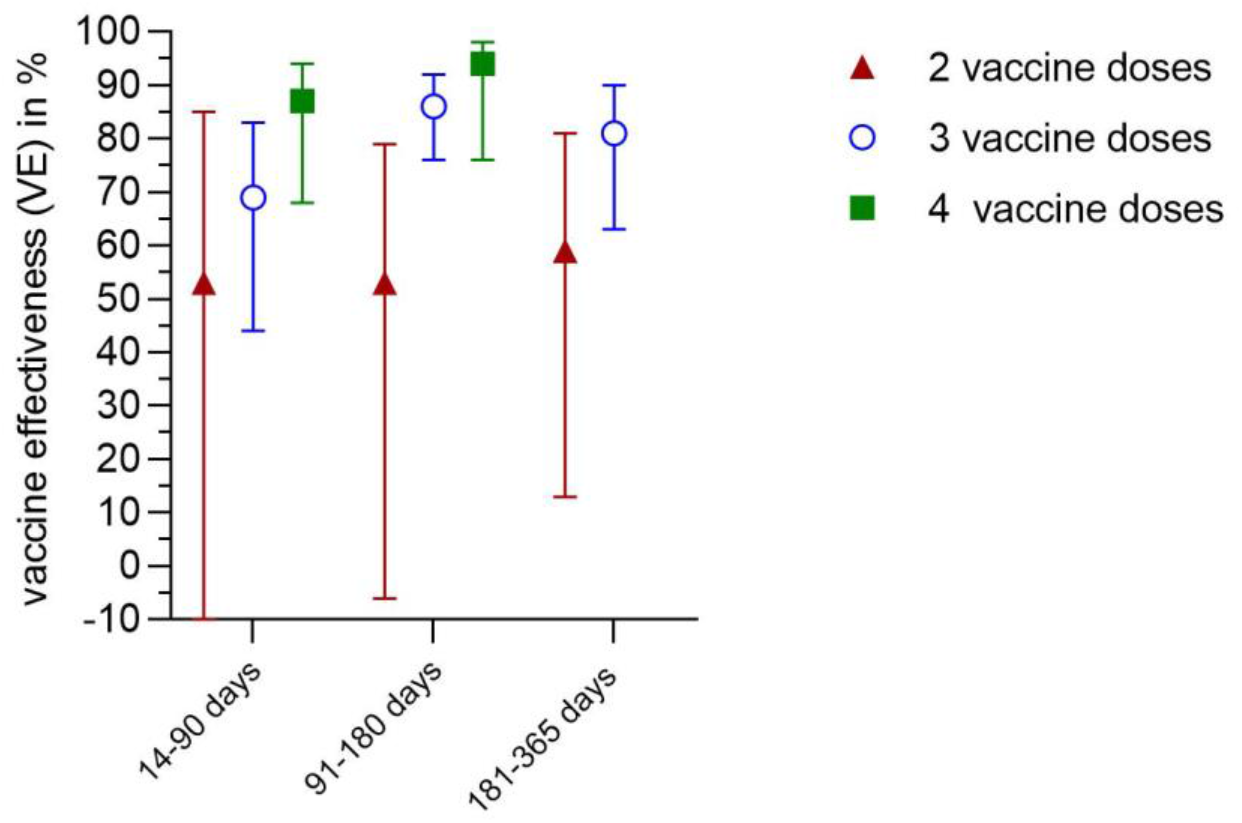
Time between last vaccine dose and symptom onset: Vaccine effectiveness for different time intervals.

## Discussion

To our knowledge, this is the first study in Germany to assess the VE of COVID-19 vaccines against severe disease caused by the Omicron variant in the post-marketing phase. Our analysis suggests that three doses of COVID-19 vaccines are highly effective in preventing hospitalization due to COVID-19 and that protection remains stable for up to one year. A fourth dose further increases the protective effect.

The proportion of older patients and cases with immunocompromising comorbidities in the Omicron wave was higher compared with our data from an earlier recruitment phase in the Delta wave [9]. This difference in clinical characteristics was confirmed in other studies [18, 19]. In settings with a high immunization coverage and a highly transmissible variant such as Omicron, immune-compromised and older persons were more often affected by severe COVID-19. Patients with comorbidities often have a fragile health situation, and even a mild or moderate SARS-CoV-2 infection can impair the state of health and lead to hospital admission with COVID-19 as the main diagnosis. However, the lower pathogenicity of the Omicron variant and the advanced therapeutical possibilities still led to milder disease in the Omicron wave compared with the Delta wave [9, 18-20].

The proportion of unvaccinated cases and controls was markedly lower in the Omicron wave compared to the Delta wave [9] and the percentage of boostered cases and controls increased. The increase might be related to the awareness campaign for vaccination in Germany and to increasing confidence in the vaccines. Nevertheless, as only 8% of the cases with an immunocompromising comorbidity had received a fourth vaccine dose, the vaccination coverage of patients at risk for severe COVID-19 can still be increased.

Whereas two vaccine doses reduced the likelihood of severe COVID-19 by 55% (95% CI: 12-78%), the effectiveness was higher for three (81%, 95% CI 68-90%) and especially four doses (97% after adjustment, 95% CI: 88-99%). In the analysis of our data from the Delta wave VE was higher for two doses (89%, 95% CI: 84-92%) and three doses (97%, 95% CI 95-99%). The results from the Omicron wave analyses were comparable with the results of other studies and systematic reviews [21-25].

A systematic review from Shao et al. including 113 studies found a lower pooled VE for Omicron (two doses 56% (95% CI:51-61) and three doses 83% (95%CI: 81-86%) compared to the VE against the Delta variant (85% (82-87%) and 93% (89-95%), respectively) for COVID-19 related hospitalization [24].

A retrospective US-based study that examined the effectiveness of BNT162b2 based on medical records from December 2021 until January 2022 reported a VE of 68% (95% CI: 58-75%) for two vaccine doses and 89% (95% CI: 84-92%) for three vaccine doses against hospital admission with COVID-19 symptoms and verified Omicron infection. No waning was observed for up to three months. The VE against the Omicron variant was lower than against the Delta variant (88% (71-95%) for two doses, 93% (89-96%) for three doses), too [21].

Šmíd et al reported a VE of 45% (95% CI: 29%-57%) against hospitalization following a positive SARS-CoV-2 test after two doses, which increased to 86% (95% CI: 84-88%) with a recent booster [23]. Since severity of infection was not verified, it is possible that a proportion of the participants had an asymptomatic or mild SARS-CoV-2 infection and were admitted for other reasons.

Ferdinands et al. [22] performed a retrospective test-negative case-control study with data from six VISION Network study sites in the US. After the second dose, the effectiveness against SARS-CoV2-associated hospitalization was 54% to 71% depending on the time lag between vaccination and symptom onset. Among recipients of three vaccine doses, VE against COVID-19 associated hospitalizations declined from 91% to 78% (after two and after more than 4 months respectively).

When Omicron was predominant, VE estimates for three vaccine doses were around 80% in our study. However, effectiveness was restored by a fourth vaccine dose (VE 96%). It must be taken into account that the fourth dose was only administered to patients at risk (over 70 years or with comorbidities) in the recruitment phase, meaning that patients with four-doses were a highly selected and rather small group.

Our analysis revealed stable protection up to one year (for three vaccine doses). As other studies showed that protection against symptomatic infection decreases noticeably with increasing time lag after the last vaccination dose [22, 23, 25-27], this is an important finding.

Additionally, our subgroup analyses (different age groups, sex, with low and high comorbidity burden, with and without ICU treatment) reveled a comparably high protection for all subgroups with VE between 75 and 92% for three vaccine doses (Supplement Table 1).

One of the main advantages of our study design is the prospective collection of detailed high-quality patient data. This permitted flexible adaptation to the different pandemic phases when the circumstances made adaptation unavoidable. To ensure high data quality, each COVID-19 diagnosis was confirmed by clinical records and - where necessary - by direct consultation of the attending physician. Only patients requiring hospitalization due to COVID-19 were included. Many post-marketing studies rely on clinical data registries, allowing a fast analysis of large datasets. However, especially in the Omicron wave, register studies have limitations. Studies based on ICD-10 code assessment can be biased through misclassification, as the coding is designed to document billing. The main diagnosis of patients in register studies may not always be COVID-19 and especially hospitalized patients with underlying comorbidities can be misclassified as severe COVID-19 cases when COVID-19 was not the disease leading to hospitalization. Thereby, VE against severe COVID-19 is likely to be underestimated in these studies, as VE against mild COVID is known to be lower.

Our study is one among few to adjust for risk of infection. The minimal set of variables for adjustment was determined by constructing a DAG (directed acyclic graph). The variable “infection risk” was necessary to build the DAG and obtain a minimal sufficient adjustment set. However, a proper assessment is not trivial since factors which influence risk of infection change over time.

The comparison of three different methods of analysis showed the robustness of the results: the results of the pairwise matched method were very similar to those of the non-matched analysis, but - as expected - not as precise [16]. For the pairwise matched analysis, a large dataset is necessary to permit analysis of subgroups.

Our study has some limitations. As selection biases are one of the major causes of bias in case-control studies we explain our approach to reduce the risk of bias and especially considerations concerning selection bias in the following. Selection biases in case-control studies assessing VE occur especially in influenza studies with outpatients, as the patient characteristics (e. g. socio-economic status) influence patients in their decision to visit a doctor or not. For this reason, test-negative design studies are preferred in this setting as they include people with a comparable health-seeking behaviour.

In contrast to this, the endpoint of our study (hospital admission) is less vulnerable towards a selection bias, as the decision of hospital admission is not strongly associated with socio-economic status and related characteristics.

In addition, we expected a selected patient group (e. g. bedridden patients with endogenous pneumonia) of test-negative patients in the pandemic as most respiratory infections were prevented by non-pharmaceutical interventions. We therefore decided against a test-negative design.

A major concern in case-control studies is selection bias. Patients who were not admitted to the hospital despite severe disease, patients with fulminant course of disease or disoriented patients who could not sign the informed consent and did not have a legal representative could not be recruited. Patients who refuse vaccination often refuse to participate in studies, too. This might lead to a selection bias at a vaccine coverage greater than 85-90% [28, 29]. Furthermore, unvaccinated people in a population with high vaccination coverage may also have a different risk compared to the general population as these patients might not adhere to non-pharmaceutical interventions. We took countermeasures in distributing incentives to participants and offered training courses to study nurses to support participant recruitment.

The second advantage of the test-negative design is reducing selection bias with regard to exposure to the virus. As the prevalence was high in the recruitment phase and we adjusted additionally for seven factors related to the risk of infection (e. g. profession, daily activity without mask, housing situation; details in the supplement), we hypothesize a low risk of bias in this respect.

Previous SARS-CoV-2 infection was not an exclusion criterion as this would have led to a selection of patients who had presumably fewer contacts, e. g. high-risk patients. SARS-CoV-2 prevalence was high during recruitment and the exclusion of recovered controls may have led to overestimation of VE as recovered patients have a lower likelihood of vaccination. However, studies comparing controls with or without previous infection reported similar results [30].

To avoid patient selection bias by the study nurse in the hospital, the potential participants were recruited in a given order (day of the birth date). Every week, study nurses filled-in a non-responder list where reasons for non-participation were documented, irregularities were followed by site visits with quality control.

To avoid a selection of patients without language barrier, an interpreter service with specially trained interpreters was employed that enabled the translation of conversations with patients in 40 languages. In addition, the questionnaires were translated into 7 languages.

As controls were usually matched in the same hospital, bias due to regional differences could be avoided. Since elective patients might be more careful with personal contacts before the hospital admission, we instructed the study nurses to preferably recruit non-elective patients.

## Conclusion

High vaccination coverage, hybrid-immunity and evolution of viral variants changed the spectrum of patients with COVID-19 in the hospitals. Patients in the Omicron wave were significantly older and had more comorbidities compared to those in the Delta wave. Nevertheless, the course of disease was milder. COVID-19 vaccines were altogether highly protective against hospitalization in real-world settings in Germany during Omicron-variant predominance. High VE was also observed in subgroup analyses and remained stable for up to one year. The results were robust in the matched as well as unmatched analysis. However, the VE was lower compared to the Delta wave, which can be explained by partial immune evasion of the Omicron variant. In further analyses, we will investigate the impact of vaccination to prevent long COVID among our study participants.

## Data Availability

All data produced in the present study are available upon reasonable request to the authors

## Acknowledgments

The authors thank all study nurses for the valuable contribution, namely Sawsanh Al-Ogaidi, Nancy Beetz, Belgin Esen, Rola Khalife, Katja Lange, Luise Mauer, Antje Micheel, Marlies Schmidt, Yvonne Weis, Franziska Weiser, Aysete Yencilek. We thank Vanessa Piechotta for critical reading of the manuscript, as well as Anna Meier, Swetlana Muminow, Richard Schensar, Ellen Busch, Hanna Buck, and Moritz Gehring for their support in organizing the project and Vincent Stoliaroff-Pépin for support with R.

