## Supplementary Material for "Vaccine effectiveness against severe COVID-19 during the Omicron wave in Germany: Results from the COViK study"

**Supplementary Information.**

### Supplementary Methods

Inclusion/exclusion criteria and matching: Cases had to be tested PCR-positive for SARS-CoV-2, with symptom onset no longer than 14 days before hospital admission. Only patients whose COVID-19 symptoms were too severe to be treated as outpatients were eligible, i. e. COVID-19 had to be the reason for hospitalization. If the degree of severity of COVID-19 was ambiguous, the physician in charge was consulted to decide whether COVID-19 of the potential case was severe and the patient had been hospitalized for this reason or not. Additionally, patients with severe nosocomial COVID-19 and patients with severe COVID-19 complications were included. A clinical case definition was not applied as the symptoms of COVID-19 and COVID-19 complications show a considerable range of variation and vary over time depending on the variant.

The analysis was restricted to cases infected with the Omicron variant. We therefore analyzed all patients included from 1 December 2021 until 5 September 2022 with a sequencing result (or PCR typing result) confirming the Omicron variant and all patients included from 28 January 2022 until 5 September 2022 with or without sequencing result for Omicron, as Omicron was the dominant variant (> 95%) in Germany from 28 January 2022 onwards [2]. Control patients were carefully selected to represent the general population. Controls were included if they were tested negative for SARS-CoV-2 by PCR. When feasible, controls were recruited from the same hospital (if not possible, from the same city) as the cases to ensure comparable regional incidences and protection measures for cases and controls. Patients with acute surgical diseases were chosen primarily. Previous SARS-CoV-2 infection was not an exclusion criterion, as this would have led to a selection bias (see "Reduction of risk of bias" in discussion). Cases and controls with contraindications for vaccination, e. g. anaphylactic reaction in the anamnesis after vaccination, or unable to sign the informed consent were not eligible to participate. Two controls were matched to each case, based on hospital admission date (+/- 14 days), age (+/- 10 years) and sex.

Biological samples: Samples were stored at 2-8°C and sent by post with attached cold packs or courier twice a week to the laboratory. All laboratory analyses were performed at the Centre for Biological Threats and Special Pathogens at the Robert Koch-Institute in Berlin. SARS-CoV-2 real-time PCR was performed to confirm or exclude a current SARS-CoV-2 infection [31]. If virus detection by PCR from the naso-/oropharyngeal swap was unsuccessful, sputum samples were requested for PCR analysis, because the sensitivity can be higher in sputum samples than in naso-/oropharyngeal swabs in the course of disease. If the PCR test failed, results of the hospital were used if available.

The virus variant was determined by sequencing using the AmpliCoV protocol [32], or by PCR typing assays if RNA load was too low for sequencing. Again, results from the hospital were used if we sequencing results were not available from the laboratory of the Robert Koch-Institute.

In addition, blood samples from cases and controls were taken, usually on the day of study inclusion. Antibodies against SARS-CoV-2 and other coronaviruses were determined by an in-house established multiplex suspension assay [33] and virus-neutralization tests were performed for future immunological analyses.

Statistical analysis: The adjustment variables are based on the DAG, Supplement Figure 2.

We adjusted for the minimal adjustment set with 10 variables:

- Age: we adjusted for three categories, 18-59 years, 60-69 years and 70-79 years. We decided for categories instead of age as a continuous variable in terms of applicability
- socio-economic status: we chose education as a proxy (see also "Discussion, School education (5.3.)) and adjusted for four categories, no graduation, 9 school years, 10 school years (secondary school certificate), 12 or 13 school years (high school graduation).
- pre-existing comorbidities: we adjusted for the number of pre-existing comorbidities
- profession: we adjusted for the group "profession with close contact to people" (medical staff with patient contact, service provider with close contact to customers, e.g. nail spa or pedicure, and social profession with close contact, e.g. teachers, educators, social workers)
- housing situation: number of household members younger than 25 years
- contact to SARS-CoV-2-positive persons in the 14 days before symptom onset (controls: before hospital admission)
- self-assessment of risk of infection (estimated probability of SARS-CoV-2-infection before symptom onset: very likely / likely or unlikely / very unlikely)
- compliance with non-pharmaceutical interventions (self-assessment: very good / good compliance or poor/no compliance))
- close contact with persons who have a high risk for severe course of COVID-19
- number of daily "high risk" activities, e.g. shopping, restaurant, choir, without mask in the 14 days before symptom onset (controls: before hospital admission). More than 2 activities 14 days before symptom onset or less than 2 activities 14 days before symptom onset

**Figures and Tables**

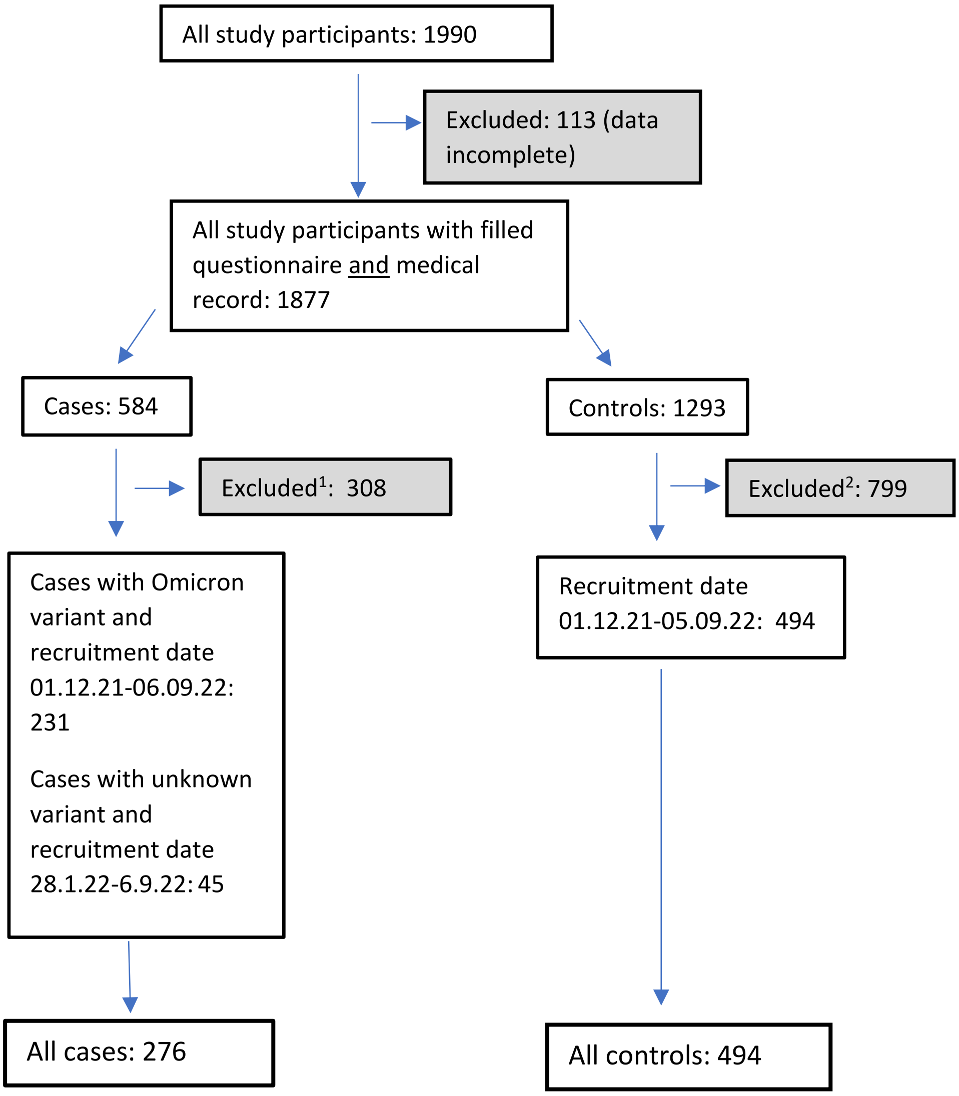


1 - recruitment before 01.12.2021 or after the 05.09.22
- or cases with Delta variant
- or cases recruited 1.12.22-27.1.22 with missing information about the variant

2 - recruitment before 01.12.2021 or after the 05.09.22

***Supplement Figure A 1 Inclusion /exclusion criteria***

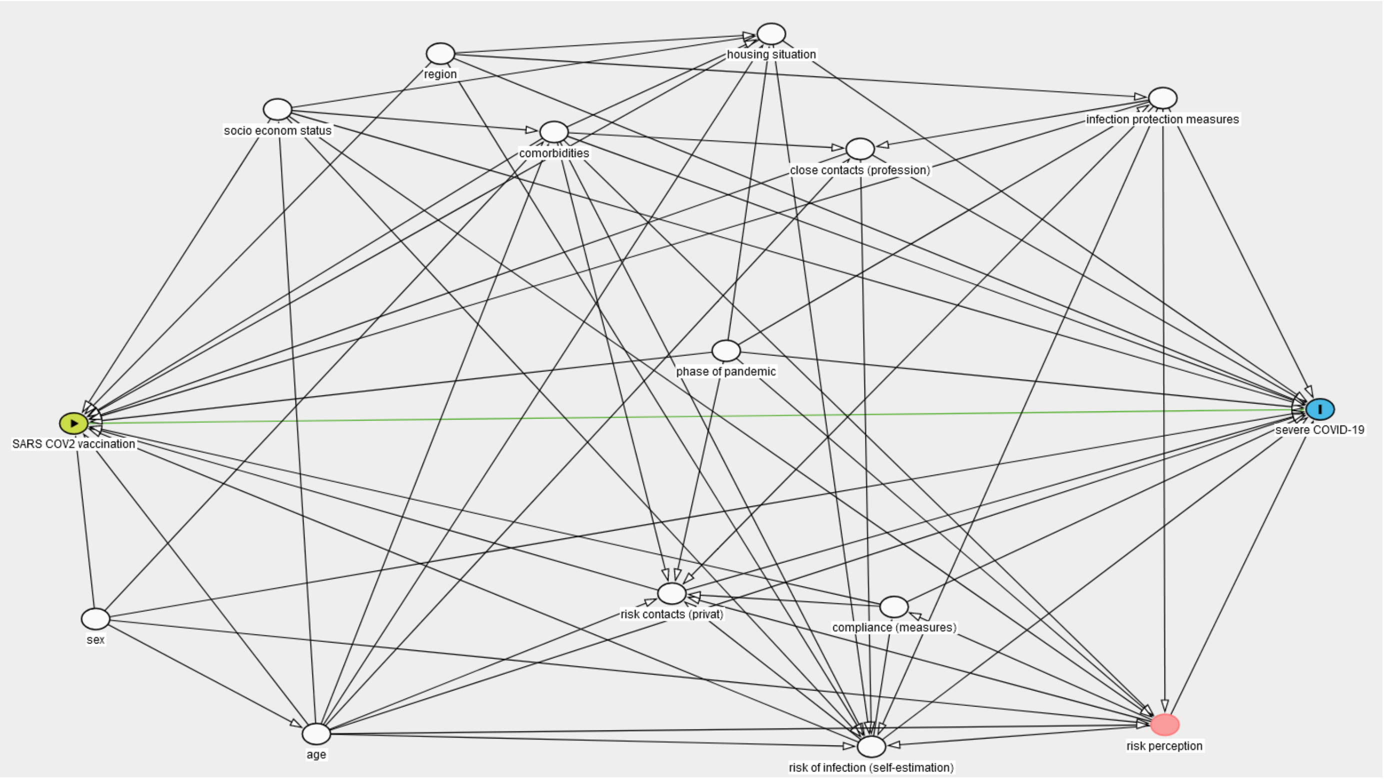


***Supplement Figure A 2 Directed Acyclic Graph (DAG) for the effect of SARS-CoV-2 vaccination (exposure) on severe COVID-19 (outcome)***

Green: exposure, blue: outcome, red: ancestor of exposure and outcome, adjustment not necessary, not adjusted, white: variable, adjusted

| | 2-dose VE (vs 0 doses) 102 cases and 75 controls | | | 3-dose VE (vs 0 doses) 201 cases and 377 controls | | |
|---|---|---|---|---|---|---|
| | Vaccinated cases n (%) | Vaccinated controls n (%) | VE (%) (95% CI) | Vaccinated cases n (%) | Vaccinated controls n (%) | VE (%) (95% CI) |
| **All patients** | 46/102 (45%) | 49/75 (60%) | 56.4 (19-76) | 145/201 (72%) | 351/377 (93%) | 80.8 (68-88) |
| **Sex** | | | | | | |
| Male | 23/48 (48%) | 31/46 (67%) | 55.4 (-2-80) | 85/110 (77%) | 205/220 (93%) | 75.1 (50-87) |
| Female | 23/54 (43%) | 18/29 (62%) | 54.6 (-14-82) | 60/91 (66%) | 146/157 (93%) | 85.4 (69-93) |
| **Age (years)** | | | | | | |
| 18-59 | 22/39 (56%) | 17/33 (51%) | -21.7 (-208- 51) | 28/45 (62%) | 112/128 (88%) | 76.4 (47-89) |
| 60-69 | 7/19 (37%) | 15/18 (83%) | 88.3 (44-97) | 32/44 (73%) | 89/92 (97%) | 91.0 (66-97) |
| 70-90 | 17/44 (37%) | 17/24 (71%) | 74.1 (24-91) | 85/112 (76%) | 150/157 (96%) | 85.3 (64-93) |
| **Pre-existing comorbidities[a]** | | | | | | |
| < 3 | 26/62 (42%) | 34/53 (64%) | 59.6 (14-81) | 85/121 (70%) | 255/274 (93%) | 82.4 (67-90) |
| ≥ 3 | 20/40 (50%) | 15/22 (68%) | 53.3 (-38-84) | 60/80 (75%) | 96/103 (93%) | 78.1 (45-91) |
| **Pre-existing comorbidities[b] of the immune system** | | | | | | |
| none | 31/68 (46%) | 41/65 (63%) | 50.9 (1-75) | 105/142 (74%) | 292/316 (92%) | 76.6 (59-86) |
| ≥ 1 | 15/43 (35%) | 8/10 (80%) | 80.2 (-7--96) | 40/59 (68%) | 59/61 (92%) | 92.8 (67-98) |
| **Time between last vaccine dose and symptom onset[c] (days)** | | | | | | |
| 14-90 | 7/63 (11%) | 7/33 (21%) | 53.3 (-46-85) | 46/102 (45%) | 70/96 (73%) | 69.4 (44-83) |
| 91-180 | 16/72 (22%) | 16/42 (38%) | 53.5 (-6-79) | 62/118 (53%) | 211/237 (89%) | 86.3 (76-92) |
| 181-365 | 20/76 (26%) | 23/49 (47%) | 59.6 (13-81) | 24/80 (30%) | 59/85 (69%) | 81.1 (63-90) |
| **ICU admission[d]** | | | | | | |
| Yes | 6/12 (50%) | 49/75 (65%) | 46.9 (-81-84) | 10/16 (63%) | 351/377 (93%) | 87.6 (63-95) |
| No | 40/90 (44%) | 49/75 (65%) | 57.5 (20-77) | 135/185 (73%) | 351/377 (93%) | 80.0 (66-88) |

Hospitalization VE: COVID-19 vaccine effectiveness against hospitalization; 95% CI: 95% confidence interval; ICU: Intensive Care Unit; NA: Not Applicable; [a] Examples of pre-existing comorbidities: heart failure, renal insufficiency, diabetes mellitus, COPD, stroke; [b] comorbidities of the immune system, e.g. cancer, HIV and patients with immune suppressive medication (e.g. after organ transplantation); [c] for controls: time between the last vaccine dose and hospital admission; [d]cases with/without ICU treatment versus controls without ICU admission treatment

***Supplement Table A 1 COVID-19 vaccine effectiveness (VE) against hospitalization after two or three vs. zero vaccine doses, non-matched, unadjusted, interim analysis of the COViK study, Omicron wave, December 2021 - September 2022, Germany***

| | 4-dose VE (vs 0 doses) 71 cases and 88 controls | | | 2 vs ≥ 3 doses 206 cases and 462 controls | | |
|---|---|---|---|---|---|---|
| | Vaccinated cases n (%) | Vaccinated controls n (%) | VE (%) (95% CI) | Vaccinated cases n (%) | Vaccinated controls n (%) | VE (%) (95% CI) |
| **All patients** | 15/71 (21%) | 62/88 (70%) | 88.7 (76-94) | 160/206 (78%) | 413/462 (89%) | 58.7 (35-73) |
| **Sex** | | | | | | |
| Male | 11/36 (31%) | 36/51 (71%) | 81.6 (53-92) | 96/119 (81%) | 241/272 (89%) | 46.3 (3-70) |
| Female | 4/35 (11%) | 26/37 (70%) | 94.5 (80-98) | 64/87 (74%) | 172/190 (91%) | 70.8 (42-85) |
| **Age (years)** | | | | | | |
| 18-59 | 1/18 (6%) | 0/16 e | - | 29/51 (57%) | 112/129 (87%) | 79.9 (57-90) |
| 60-69 | 3/15 (20%) | 6/9 (67%) | 87.5 (18-98) | 35/42 (83%) | 95/110 (86%) | 21.0 (-109-70) |
| 70-90 | 11/38 (29%) | 56/63 (89%) | 94.9 (85-98) | 96/113 (85%) | 206/223 (92%) | 53.3 (4-77) |
| **Pre-existing comorbidities[a]** | | | | | | |
| < 3 | 7/43 (16%) | 38/57 (67%) | 90.2 (74-96) | 92/118 (78%) | 293/327 (90%) | 58.9 (27-76) |
| ≥ 3 | 8/28 (29%) | 24/31 (77%) | 88.3 (62-96) | 68/88 (77%) | 120/135 (89%) | 57.5 (11-79) |
| **Pre-existing comorbidities[b] of the immune system** | | | | | | |
| none | 8/45 (18%) | 50/74 (68%) | 89.6 (74-95) | 113/144 (78%) | 342/383 (89%) | 56.3 (27-73) |
| ≥ 1 | 7/26 (27%) | 12/14 (86%) | 93.8 (65-98) | 47/62 (76%) | 71/79 (90%) | 64.6 (10-86) |
| **Time between last vaccine dose and symptom onset[c] (days)** | | | | | | |
| 14-90 | 8/64 (13%) | 29/55 (53%) | 87.1 (68- 94) | 54/61 (89%) | 99/106 (93%) | 45.4 (-63-81) |
| 91-180 | 2/58 (3%) | 18/44 (41%) | 94.8 (76-98) | 64/80 (80%) | 229/245 (93%) | 72.0 (41-86) |
| 181-365 | 3/29 (10%) | 2/58 (3%) | 69.0 (-96- 95) | 26/46 (57%) | 62/85 (73%) | 51.7 (-2-77) |
| **ICU admission[d]** | | | | | | |
| Yes | 0/6[e] | 62/88 (70%) | 96.5 (35-99) | 10/16 (63%) | 412/461 (89%) | 80.2 (43-93) |
| No | 15/65 (23%) | 62/88 (70%) | 87.4 (73-93) | 149/189 (79%) | 412/461 (89%) | 55.7 (30-72) |

Hospitalization VE: COVID-19 vaccine effectiveness against hospitalization; 95% CI: 95% confidence interval; ICU: Intensive Care Unit; [a] Examples of pre-existing comorbidities: heart failure, renal insufficiency, diabetes mellitus, COPD, stroke; [b] comorbidities of the immune system, e. g. cancer, HIV and patients with immune suppressive medication (e. g. after organ transplantation); [c] for controls: time between the last vaccine dose and hospital admission; [d]cases with/without ICU treatment versus controls without ICU admission treatment; [e] calculated with 0.5/6.

***Supplement Table A 2 COVID-19 vaccine effectiveness (VE) against hospitalization after four vs. zero and two vs. ≥ three vaccine doses, non-matched, unadjusted, interim analysis of the COViK study, Omicron wave, December 2021 - September 2022, Germany***

| | VE (%) after 3 doses (95% CI) | VE (%)after 4 doses (95% CI) |
|---|---|---|
| Non-matched analysis | 80.8% (68-88%) | 88.7% (76; 94%) |
| Matched pair analysis | Control 1: 82.5% (64-92%)<br>Control 2: 86.5 %(69-94%) | Control 1: 83.3% (-1231-99%)<br>Control 2: 90.0% ( -106-99%) |

***Supplement Table A 3 Results of pairwise matched analysis vs. non-matched analysis: vaccine effectiveness (without stratification)***

| | Non-matched analysis | Matched analysis |
|---|---|---|
| | VE (%) after 3 doses (95% CI) | VE (%) after 3 doses (95% CI) |
| male | 75.1% (50-87%) | 85.7% (57-95%) |
| female | 85.4% (69-93%) | 84.6 % (60-94%) |
| 18-59 years | 76.4% (47-89%) | 77.8% (-13-96%) |
| 60-69 years | 91.0% (66-97%) | 94.4% (43-99%) |
| 70-90 years | 85.3% (64-93%) | 81.0% (47-93%) |
| < 3 comorbidities[a] | 82.4% (67-90%) | 87.5% (64-96%) |
| ≥ 3 comorbidities[a] | 78.1% (45-91%) | 77.8% (-13-96%) |

a: Examples of pre-existing comorbidities: Heart failure, renal insufficiency, diabetes mellitus, COPD, stroke

***Supplement Table A 4 Results of pairwise matched analysis vs. non-matched analysis: subgroup analysis of vaccine effectiveness for participants with three vaccine doses (versus 0)***

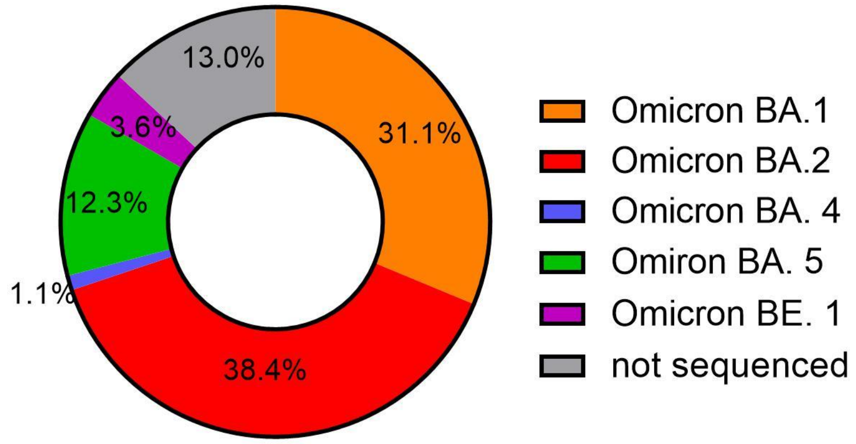


***Supplement Figure A 3 Proportions for Omicron variants***

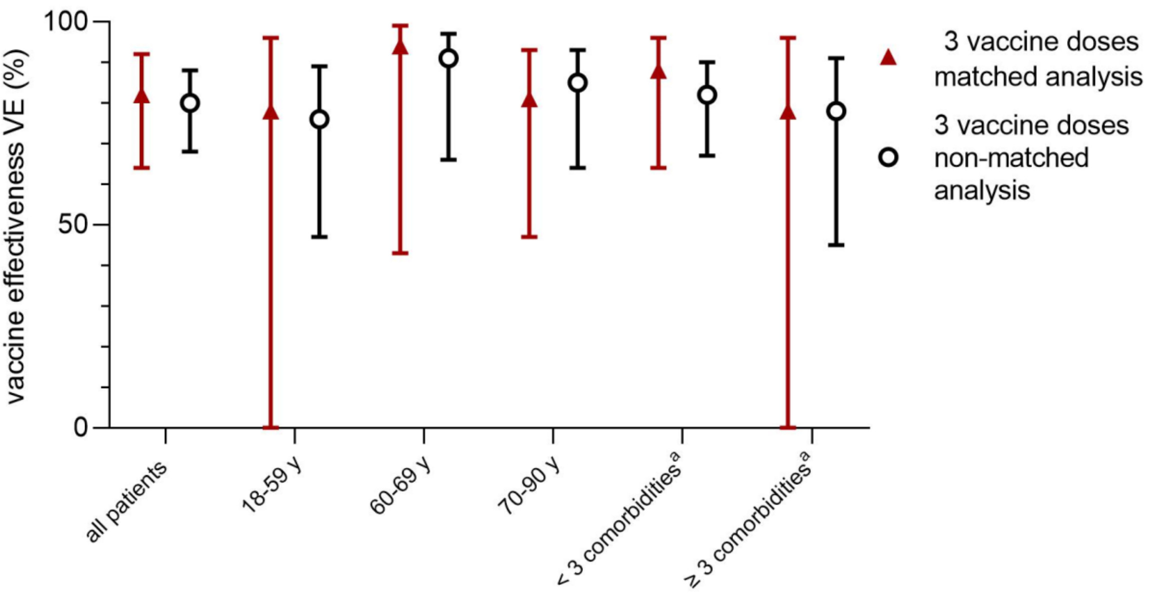


***Supplement Figure A 4 Comparison of vaccine effectiveness in the matched and non-matched analysis***
